# Geospatial characterizing of Under-Five Mortality in Alexandria, Egypt

**DOI:** 10.1101/2021.04.24.21255886

**Authors:** Mahmoud Adel Hassan, Ahmed Ramadan, Mohamed Mostafa Tahoun, Abdel Rahman Omran, Shaimaa Gad EL Karim Ebrahim Ali, Ola Fahmy Khedr Esmail, Ehab Mohamed Adel Elrewany, Passent Ehab El-din Ahmed El-Meligy, Amira Mahmoud Elzayat, Dina Hussein el Malawany, Amira Saad Mahboob, Mohamed Kamal Eldewiki, Esraa Abdellatif Mohamed Abdellatif Hammouda, Ramy Mohamed Ghazy

**Affiliations:** Institute of Graduate Studies and Research – Alexandria University; Clinical Research Unit, EVA Pharma; Epidemiology Department, High Institute of Public Health, Alexandria University; Ministry of Health and Population; Public Health Nursing, High Institute of Public Health, Alexandria University; The Egyptian Pharmacovigilance Center -Alexandria Regional Center. Egyptian Drug Authority; Tropical Health Department, High Institute of Public Health, Alexandria University; Alexandria Main University Hospital; Nutrition Department, High Institute of Public Health, Alexandria University; Environmental Health Department, High Institute of Public Health, Alexandria University; Information Professional, High Institute of Public Health, Alexandria University

**Keywords:** Geographic information system, under-five mortality, death records, Alexandria, structural equation model, hot spot areas

## Abstract

This study aimed to identify geo-spatial pattern of under-five mortality (U5M) in Alexandria and its key determinants. We analyzed the geospatial distribution of 3064 deaths registered at 24 health offices reported from January 2018 to June 2019. The localities of Alexandria city were clustered into high and low incidence areas. Neonates represented 58.7% of U5M, while post-neonates and children were 31.1%, 10.2% respectively. Male deaths were significantly higher (*P*=0.036). The main leading causes of U5M were prematurity (28.32%), pneumonia (11.01%), cardiac arrest (10.57%), congenital malformation (9.95%), and childhood cardiovascular diseases (9.20%). Spatial distribution of U5M (including the most common three causes) tend to be clustered in western parts of Alexandria (El Hawaria, Bahig, Hamlis and Ketaa Maryiut). Another 9 clusters are at risk of being hotspots. Illiteracy, divorce, and poor locality characteristics (household size, population density, and access to water supply and sanitation), were statistically significant predictors of U5M.

## Introduction

Despite the achieved progress in reducing under-five mortality (U5M) during the past few decades all over the world, it was estimated that 5.4 million children died before the age of 5 in 2017. ^(1)^ The global burden of U5M necessitates action to achieve further reduction in child mortality. Accordingly, reducing child mortality is of target 3.2 in the Sustainable Development Goals. This goal entails reducing neonatal mortality to at least as low as 12 per 1,000 live births and U5M to at least as low as 25 per 1,000 live births.^(2)^

Determinants of U5M were repeatedly considered at both community and national levels. In fact, community and environmental conditions like poor access to drinking water and sewage disposal have considerable impacts on U5M. ^(3, 4)^ Besides, some socioeconomic conditions like parents’ education level, number of under-five children (U5C) living in the same context, and child order affected the incidence of U5M. ^(5-8)^ Furthermore, maternal and child factors like child immunization and birth weight, practicing breastfeeding, birth interval, mother’s age, and contraceptive use are considered to be important determinants of U5M. ^(9)^ In hazard-prone areas, environmental risks are responsible for a part of U5M. In China, 934 children aged below 5 years died due to 21 earth quack-hit. The number of deaths is strongly associated with earthquake intensity, collapsed house, and slope. ^(10)^ Contribution of infectious diseases cannot be overlooked, pneumonia, malaria and diarrheal diseases still represent a major killer of U5C in regions vulnerable to epidemics. ^(11)^ Recently, behavioral factors such as mothers practice and perceived benefits of the latest treatment guidelines were added to the long list of the determinants of U5M. ^(11)^ At a national level, economic conditions in terms of household poverty and income per capita were suggested to be one of the key predictors for U5M as it reflects health spending. ^(12) (13)^ The low-income levels has been associated with negative impacts on access, availability, accessibility, and quality of health services and healthy diet.^(14)^

It should be noted that a few numbers of previous research considered spatial analysis of U5M clustering in Egypt. The only exception was the study undertaken by Mohamed et. al, ^(15)^ in 2004 who applied Geographical information system (GIS) in analyzing U5M in Alexandria for the period 1996-2001. For this purpose, the researchers used GIS in producing several thematic maps for incidence rate of U5M and some sociodemographic indicators. Then they visually compared the distribution pattern of high incidence rate and the considered indicators, therefore, high incidence rates revealed in some areas of Alexandria city were explained by deteriorating sociodemographic and environmental conditions in these areas. It is worth mentioning that such an analysis did not rely on a sound statistical tool and relied mainly on visual interpretation of the distribution pattern.

This paper is intended to apply GIS in mapping and analyzing the spatial pattern of U5M in Alexandria city, delineating the statistically significant hot and cold spots, and identifying the key determinants of U5M. It is thought that such an analysis and assessment that relied mainly on spatial statistics analysis tools can provide a better understanding of the key factors underlying U5M in Egypt. Consequently, this work may guide policy-makers to implement more effective intervention measures in the future.

## Data and Methods

### Study area

Alexandria city is the second-largest urban center in Egypt. Alexandria plays a crucial role in the Egyptian economy, as it is hosting the largest port in Egypt, and hosts a considerable proportion of industrial investment in Egypt. The population count of Alexandria city was estimated to be about 5.2 million in 2017 accounting for 5.4% of the total country population. ^(16)^ Alexandria city consists of 9 districts in addition to the new satellite town (New Burg El Arab town). These 9 districts are divided into 21 sections, which in turn subdivided into 143 localities (**Figure 1**).

**Figure (1):**
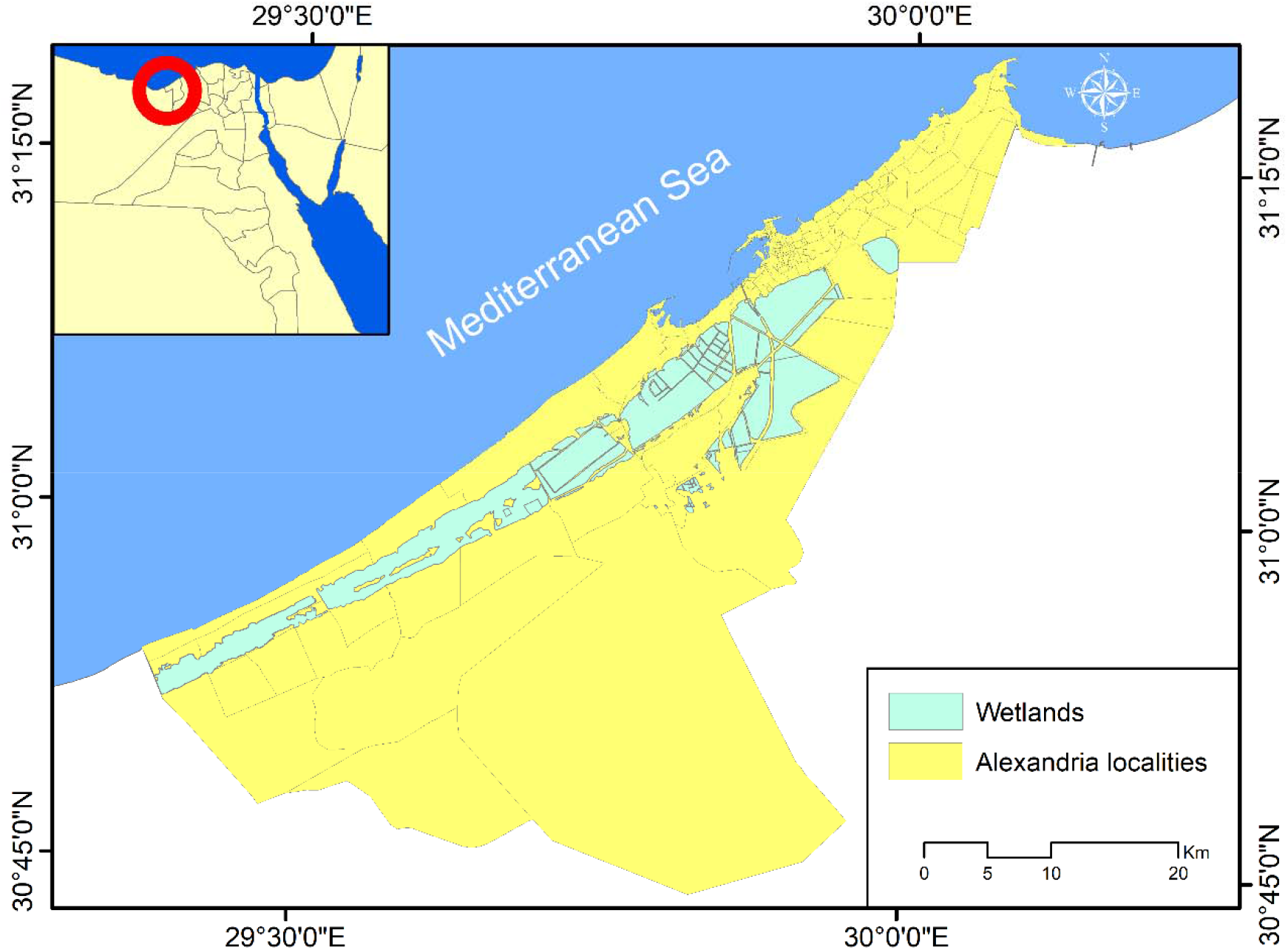
Administrative localities of Alexandria Governorate.

The population of the city has increased by about 25.2% during the period 2006-2017 ^(17)^ with an annual change rate of 2.3%, which is relatively higher than the national rate. ^(18)^ Under-five population represents about 12.1% of the total population of Alexandria. Urban population represent about 98.7% of the total population of the city, while the urban population represents 1.3%. ^(19)^

### Data

Attaining the above-mentioned objective requires access to data of U5M in Alexandria. Data was collected through a cross-sectional survey started in July to October 2019. This retrospective descriptive study was conducted in two phases: during the first phase, an inventory of U5M was developed based on registration data available at various health care offices. The survey involved collecting data of 3064 deceased children registered in 24 health offices in Alexandria during the period from 01/01/2018 to 30/06/2019. For each child, age, sex, address, date of birth and death, and the cause of death for each deceased was collected. Infant mortality included any deaths occurred before the first birthday: including neonatal deaths (death before 28 days) and post-neonatal deaths (death after 28 days and before the end of 1^st^ year). U5M included infant and child deaths (any death occurs between 1-4 years). Causes of death were coded according to the 10^th^ International Classification of Diseases version 10 (ICD 10).^(20)^

Thereafter, a geo-database for the collected data was developed, and plotted U5M through their address (**Figure 2**). During data validation, 140 deaths were excluded as they were not living in Alexandria, or they had unspecified housing address. A total of 2924 deceased children were included in the analysis.

**Figure (2):**
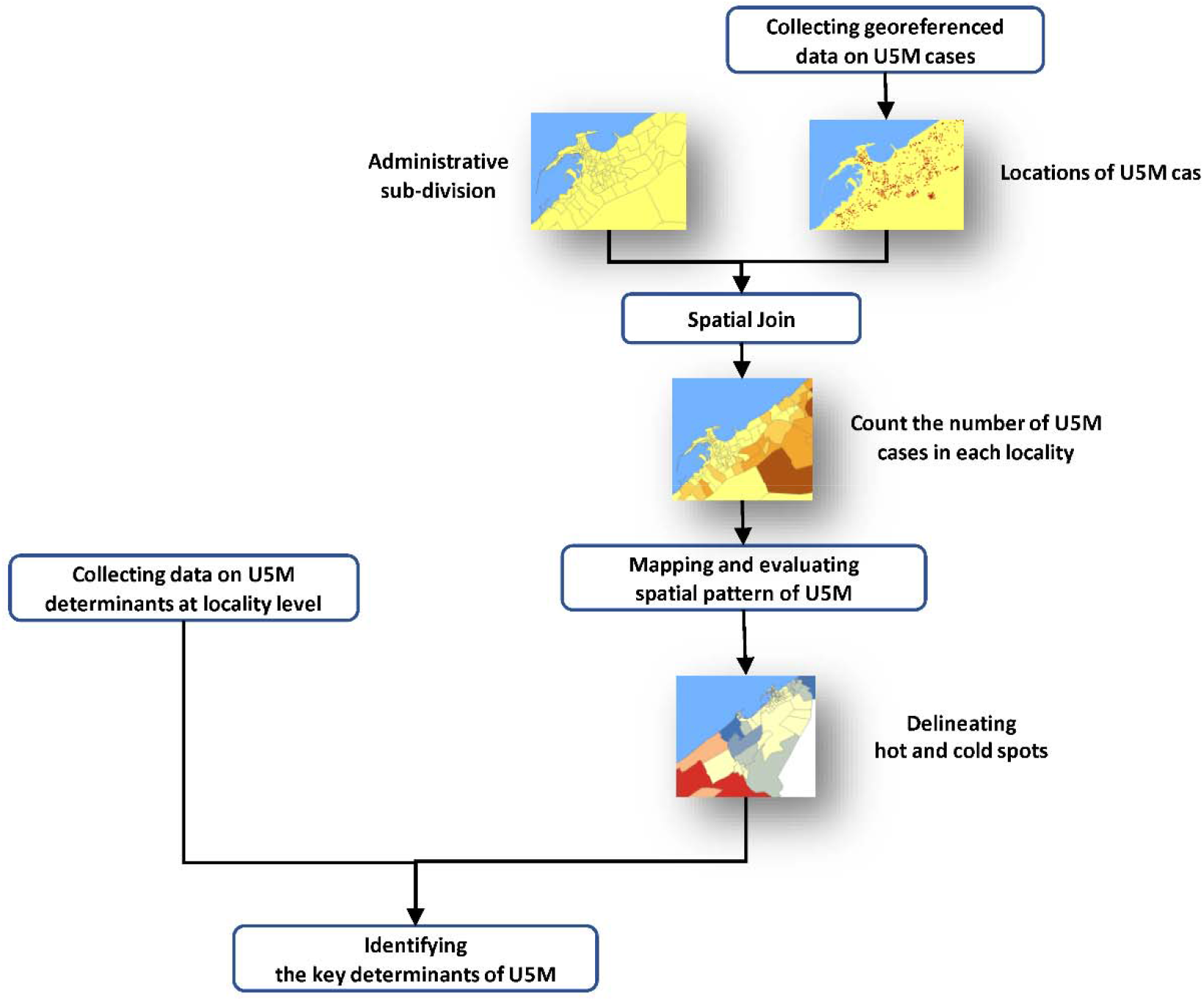
**Methodology developed for evaluating spatial pattern of under-five mortality in Alexandria using GIS**

Afterwards, a geodatabase for the collected data was developed, and data on various hypothetical sociodemographic and economic factors at locality levels were collected from the most recent population census (2017) and integrated into the developed geodatabase.

## Methods

The developed geodatabase was employed for counting the number of U5M per each locality (administrative unit) through spatial join analysis. Thereafter, the spatial pattern of U5M was examined and evaluated through Spatial Autocorrelation analysis (Moran’s I index) that highlights whether the spatial pattern is clustered, dispersed, or random. The analysis involves calculating Moran’s I Index value and both a z score and *P*-value to evaluate the significance of that Index according to the following formula:

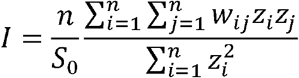

Where:

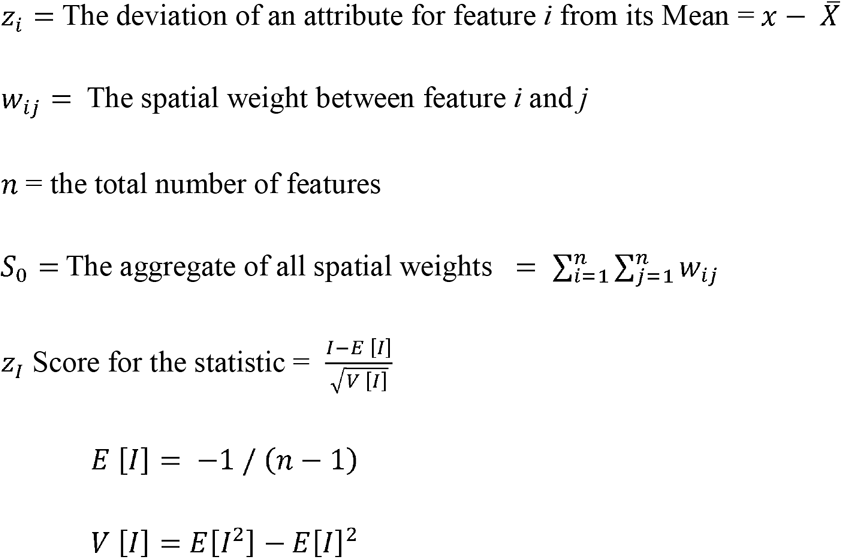

To identify the weighting scheme that was employed in calculating Moran’s I index, inverse distance method was applied, where nearby neighboring features have a larger influence on the computations for a target feature than features that are far away.

Moreover, hot spot analysis was performed to delineate those localities of statistically significant hot and cold spots in U5M. Generally, for each locality, to be a statistically significant hot spot, the locality should have a high value and be surrounded by other localities with high values as well. Hot spot analysis identifies statistically significant hot spots and cold spots through the Getis-Ord 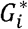 statistic for each feature (locality) in the dataset as per the following formula:

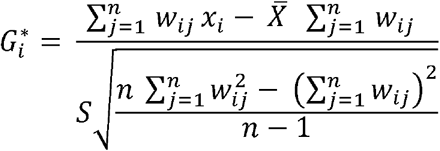

Where:

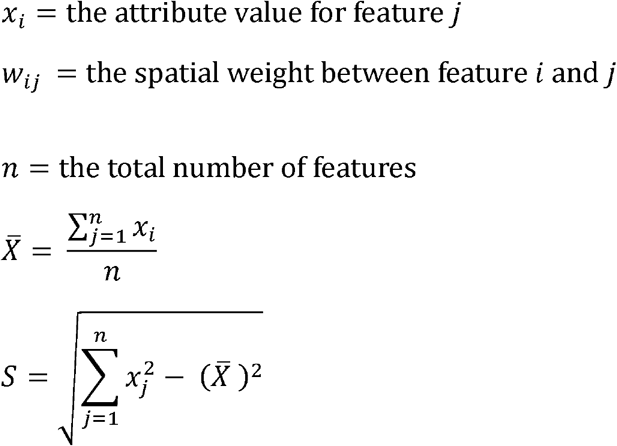

Finally, various socio-demographic characteristics of the hot and cold spot were explored to identify the key determinants of U5M in Alexandria city. Cluster analysis was done to group areas that are more like each other than others in the same group (cluster). Categorical variables were performed using chi-square test for categorical variables. The P-values of less ≤ 0.05 was considered statistically significant. Structural equation model followed; it often invokes a measurement model that defines latent variables using one or more observed variables and a structural model that imputes relationships between latent variables.

We further aimed at quantifying the interaction between the collected data (predictors) with the recorded U5M for each locality in terms of male mortality, female mortality, and total mortality records. Because of the established multi-collinearity (correlations) between the predictors, we utilized the partial least square based structural equation modelling (PLS-SEM) for the regression. The multi-collinearity issue was resolved by forming four latent variables as follows:

i) Locality characteristics consisted of the household size, access to sanitation, access to drinking water, crowding rate, and population density at each locality.

ii) Divorce, consisted of the number of divorced females, number of divorced males, and the percentage of divorced subjects at each locality.

iii) Illiteracy consisted of the illiteracy in males and females, average illiteracy, University grades for females and males, and average university grades.

iv) Dependent outcome: Mortality, consisted of U5M in males, females, and total mortalities.

### Ethical approval

Before the research was conducted, ethical clearance was sought and approved from the Ethics Committee of High Institute of Public Health, Alexandria University. For the sack of confidentiality, data sets used were anonymous of participant identity.

## Results

It was found that about 58.7% (1717) of the recorded U5M were neonatal deaths, while 31.1% (910) were post-neonates and 10.2% (297) were children aged 1-5 years. Male and female deaths represented 53.2% (1557) and 46.8% (1367) respectively. The difference between both sexes was statistically significant (X^2^ = 6.67, *P*=0.036). Table 1 This difference was evident in infant mortality; a higher proportion of males died in the neonatal period (X^2^ =5.29, *P*= 0.0214), however, there was no statistically significant difference in other age groups P □ 0.05.

**Table (1):** Deaths by age and sex among under-five children Alexandria, Egypt 2019.

| Group |  |  | Sex |  |  |  | X<sup>2</sup> | P |
| --- | --- | --- | --- | --- | --- | --- | --- | --- |
|  |  |  | Male |  | Female |  |  |  |
|  |  |  | n | % | n | % |  |  |
| Under-five<br>(2924) | Infants<br>(2627) | Neonates (1717) | 937 | 54.6 | 780 | 45.5 | 6.67 | 0.036 |
|  |  | Post-neonates (910) | 453 | 49.8 | 457 | 50.2 |  |  |
|  | Children (297) |  | 167 | 56.2 | 130 | 43.8 |  |  |

U5M: under-five mortality; Neonatal mortality vs. post neonatal ( $X^2=5.29$ , $P= 0.0214$ ); post neonatal vs. child mortality ( $X^2=3.47$ $P 0.620$ ); neonatal mortality vs. child mortality ( $X^2 =0.218$ , $P= 0.64$ ).

U5M was found to be unevenly distributed within different localities of Alexandria governorate. Generally, newly developed areas in western, southwestern, and eastern parts of the city showed the highest number of U5M compared to other parts of the city. Meanwhile, the least number of U5M were recorded in the inner old parts of the city (**Figure 3)**.

**Figure 3:**
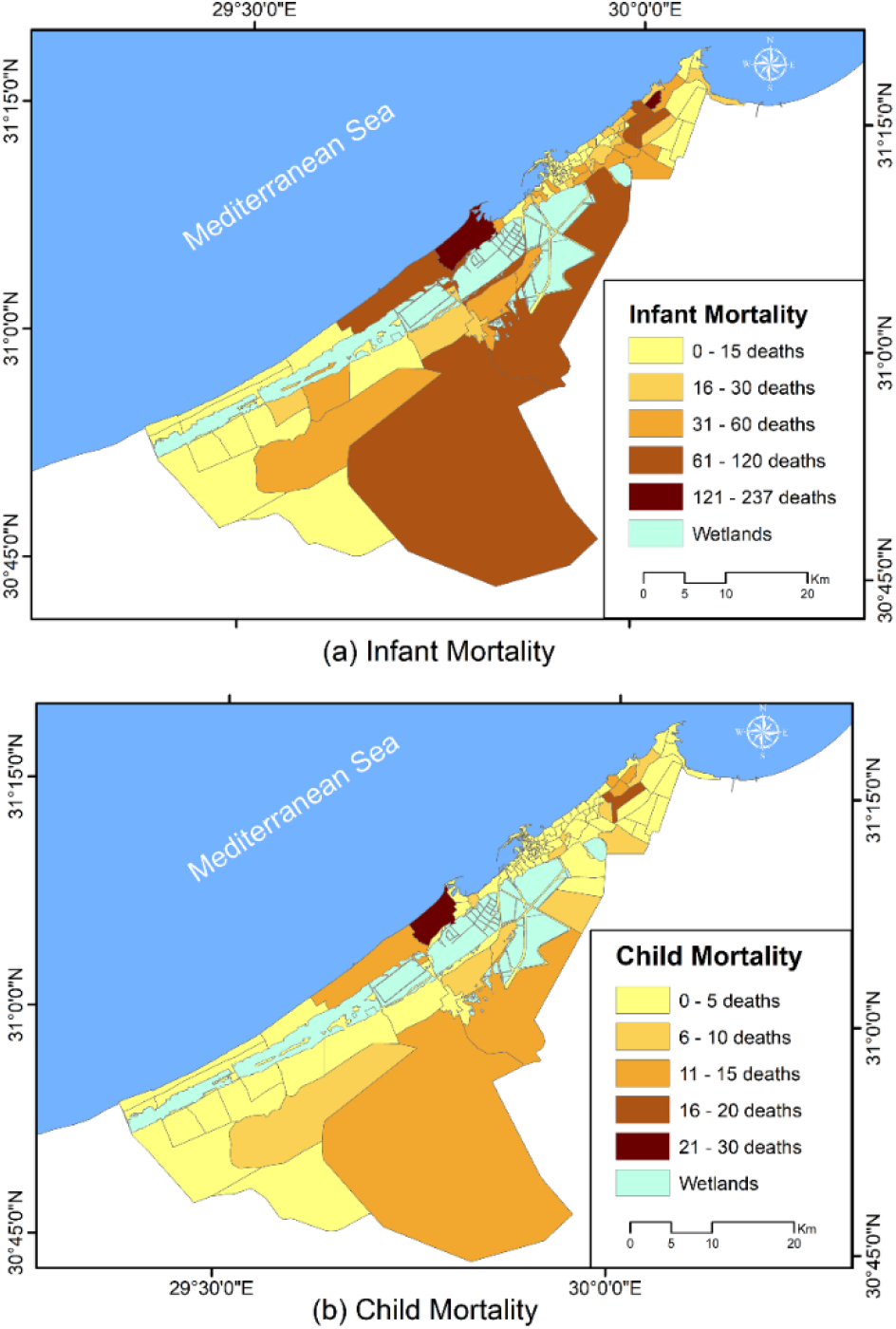
Number of under-five mortality in Alexandria by locality.

Such uneven distribution required evaluating the distribution pattern of U5M at locality level in Alexandria city. For this purpose, Spatial Autocorrelation analysis (Moran’s I index) was applied. The results of Moran’s I index revealed that there is positive spatial autocorrelation in U5M within different localities of Alexandria. The Moran’s I index recorded a small value (0.17) for U5M and high z-score recording 11.91, *P* < 0.01, respectively. Accordingly, it can be argued that, based on statistically significant *P*-value and high positive z-score, the spatial distribution of U5M tends to be clustered and this clustered pattern could not be the result of random chance. Also, the distribution of infant deaths tends to be clustered (Moran’s I index of 0.12, z-score=7.48, *P* < 0.01). Similarly, distribution of child (1-5 years) deaths was found to be clustered (Moran’s I index = 0.15, z-score =10.2, *P* < 0.1). This highlights that the clustered distribution pattern of U5M could not be the result of random chance.

Hot spot analysis was applied to delineate the main hot and cold spots for U5M in Alexandria. As a result of this analysis, a z-score for each locality in Alexandria was returned, where significant positive z-scores refer to the more intense the clustering of high values (hot spot) and significant negative z-scores indicate more intense the clustering of low values (cold spot). Generally, two cold spots were identified involving 26 localities in the north eastern and middle parts of Alexandria, while a hot spot was noticed in the western parts of the city including four localities: El Hawaria, Bahig, Hamlis and Ketaa Maryiut (**Figure 4)**

**Figure 4:**
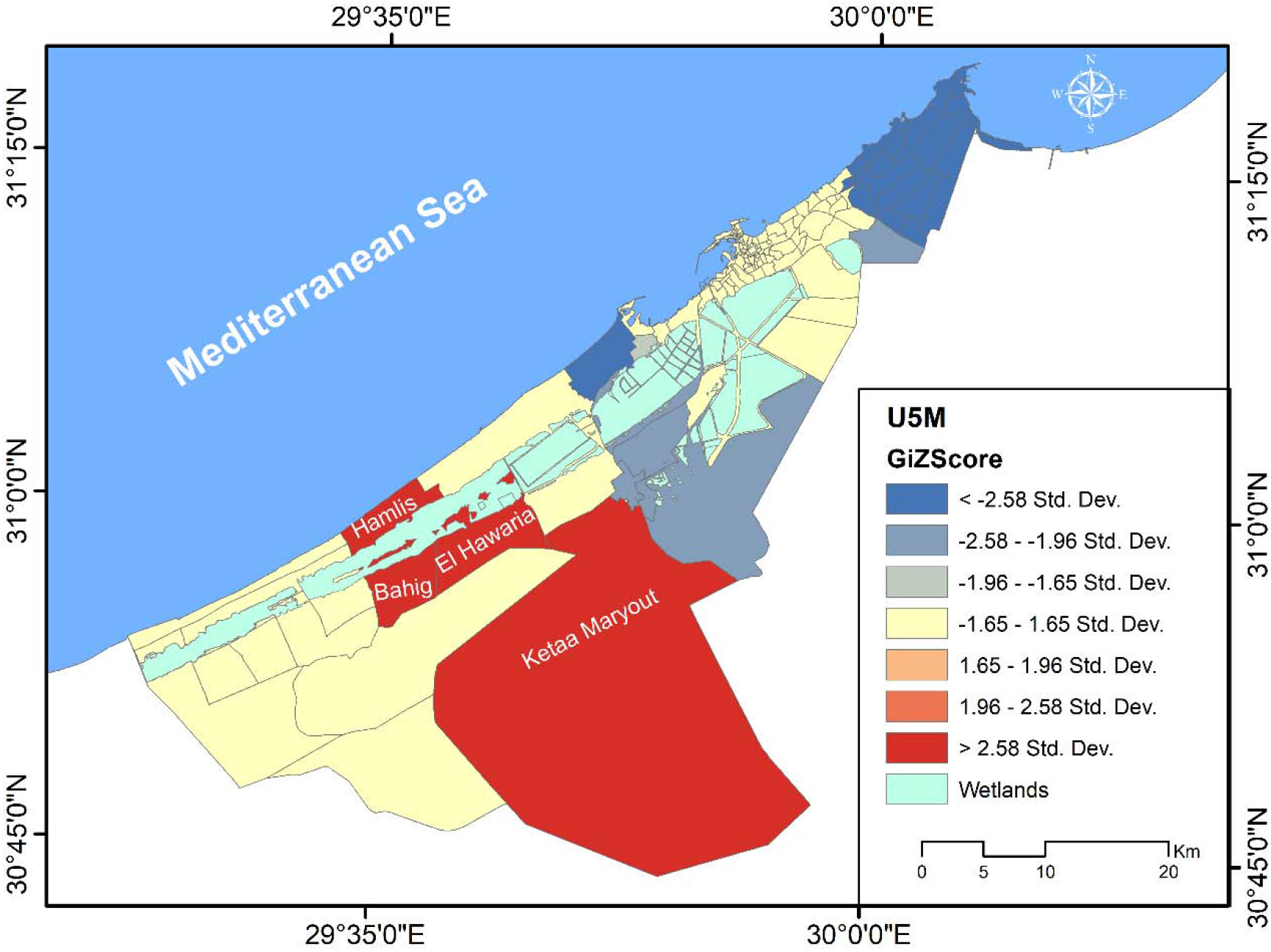
Hot spot analysis of under-five mortality Alexandria Egypt.

In an attempt to identify the main predictors of U5M in Alexandria, various potential sociodemographic determinants were explored in both cold and hot spot localities. For this purpose, data on crowding rate, household size, and educational level, access to sanitation and access to drinking water at locality level were collected from population census. Compared to cold spot localities, the hot spot localities were generally characterized by predominance of rural communities with a relatively declines sociodemographic conditions.

As for main leading causes of U5M in Alexandria, it was found that prematurity was found to be the highest contributor to U5M, where it was the leading cause for about 28.32% of the total U5M. Meanwhile, pneumonia came in the second-order as one of the leading causes of U5M with 11.01% of total deaths. Cardiac arrest and congenital malformation contributed to 10.57% and 9.95% of the total U5M. This is followed by childhood cardiovascular diseases (9.2%), septic shock (7.66%) and finally respiratory failure (4.79%) (**Figure 5**).

**Figure 5:**
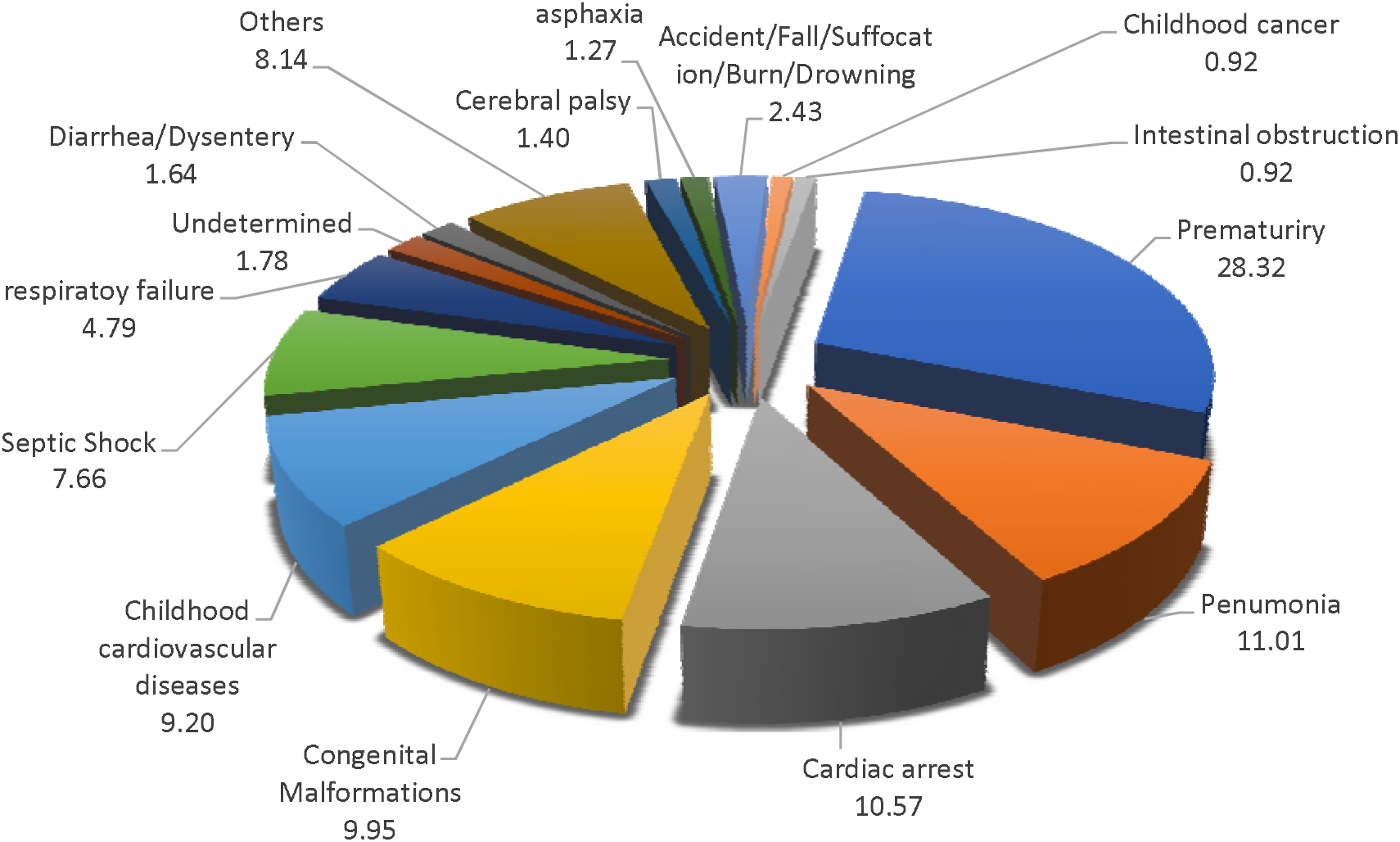
Causes of under-five mortality in Alexandria, Egypt.

In this study, the main causes of neonatal deaths were prematurity (40.36%) congenital malformations (11.18%), cardiac arrest (9.55%), congenital pneumonia (8.33%), septic shock (8.15%), and cardiovascular diseases (7.98%). Regarding causes of post-neonatal mortality pneumonia, and prematurity were the main killer (14.84% for each), followed by cardiac arrest (13.41%), childhood cardiovascular diseases (12.31%), congenital malformations (9.12%), septic shock (7.47%), respiratory failure (5.60%) and diarrheal (3.63%). Pneumonia still a main killer of children aged 1-4 years (14.97%). It is worthy to mention that accidents, drowning, and suffocation as preventable causes of childhood deaths resulted in 20.07% of the overall deaths. Cardiac arrest and cerebral palsy contributed equally (7.82%) for each, followed by septic shock (7.14%), childhood cardiovascular diseases (6.8%), and congenital malformations (5.44%). **Table 2**

**Table 2:**
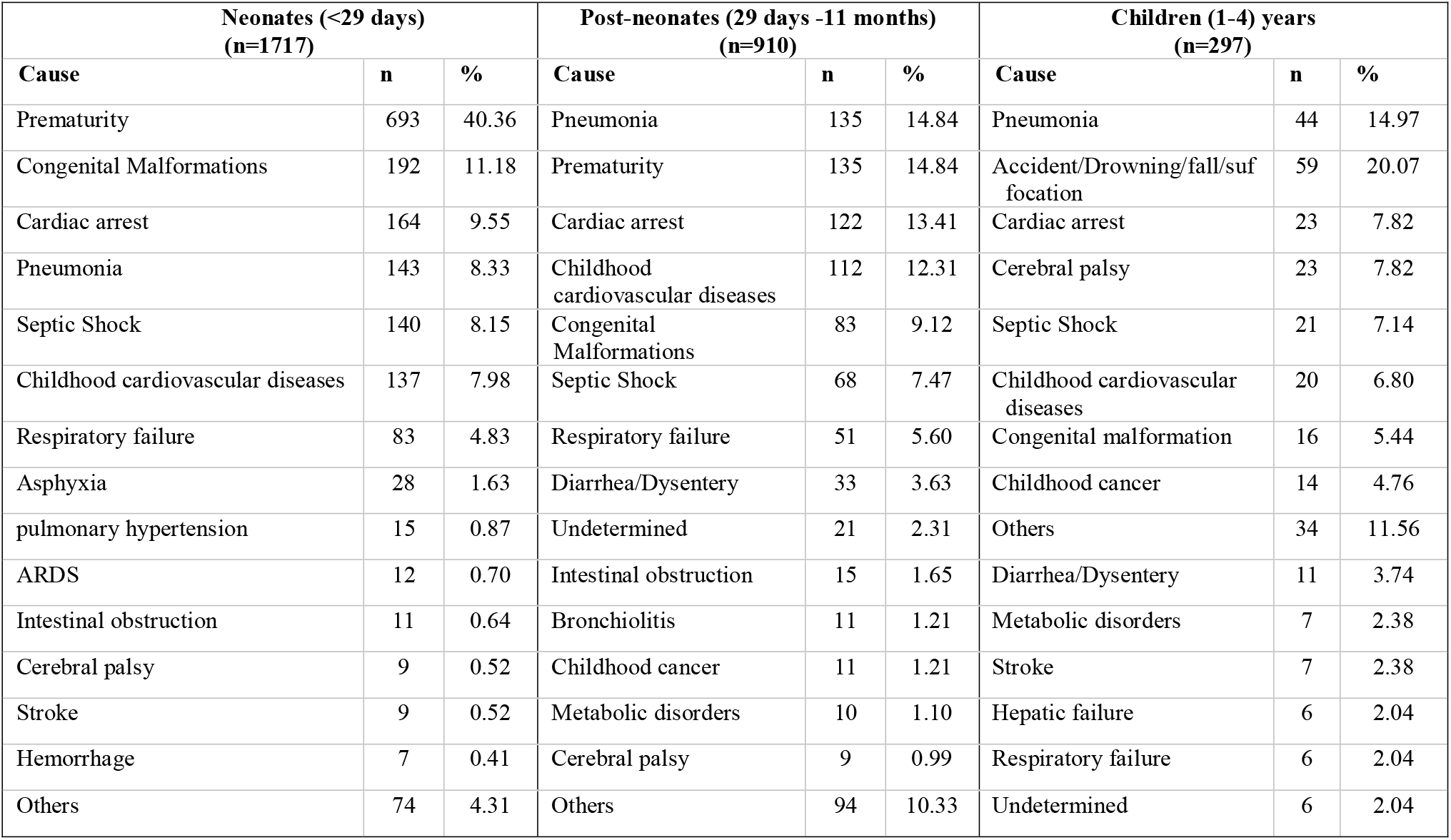
Causes of Under-five mortality by age category.

**Table 3:** Two Step Cluster analysis of localities based on collected data.

| Cluster | Cluster 1 (Mean or %) | Cluster 2 (Mean or %) |
| --- | --- | --- |
| Size | 17 localities (56.7%) | 13 localities (43.3%) |
| Classification variables |  |  |
| University grade for females | 3.56 | 22.98 |
| University grade average | 4.12 | 24.14 |
| University grade for males | 4.65 | 25.24 |
| Illiteracy average | 35.53 | 11.75 |
| Illiteracy female | 40.94 | 13.90 |
| Illiteracy male | 30.57 | 9.71 |
| Divorce in females | 44.56% | 20.34% |
| Divorce average | 22.71% | 11.74% |
| Household size | 4.16 | 3.75 |
| Population density | 7,492.06 | 50,220.08 |
| Divorce in males | 9.30% | 2.13% |
| Crowding rate | 1.21% | 1.08% |
| Access to sanitation | 66.08 | 96.98 |
| Access to drinking water | 96.72 | 98.59 |
| Status (Hot or cold spot) | 76.5% cold<br>13 cold/ 4 hot | 100% cold<br>13 cold |

\*classification inputs are ordered according to their importance for the classification

To examine the spatial distribution pattern of prematurity, pneumonia and cardiac arrest, as the main leading causes of U5M in Alexandria, Moran’s I index was applied. The results of Spatial Autocorrelation analysis (Moran’s I index) revealed small values of Moran’s I index and high z-score (*P*-value < 0.01). Such statistically significant *P*-value and high positive z-score refer to the spatial distribution of deaths of these three leading causes tend to be clustered. Therefore, hot spot analysis outlined the main cold and hots spots of these three leading causes (**Figure 6)**. However, it was noted that the distribution pattern of cold and hot spots of the three leading causes showed a certain level of similarity, where two cold spots were identified in the northeastern and middle parts of Alexandria, while a hot spot was identified in the western parts of the city.

**Figure 6:**
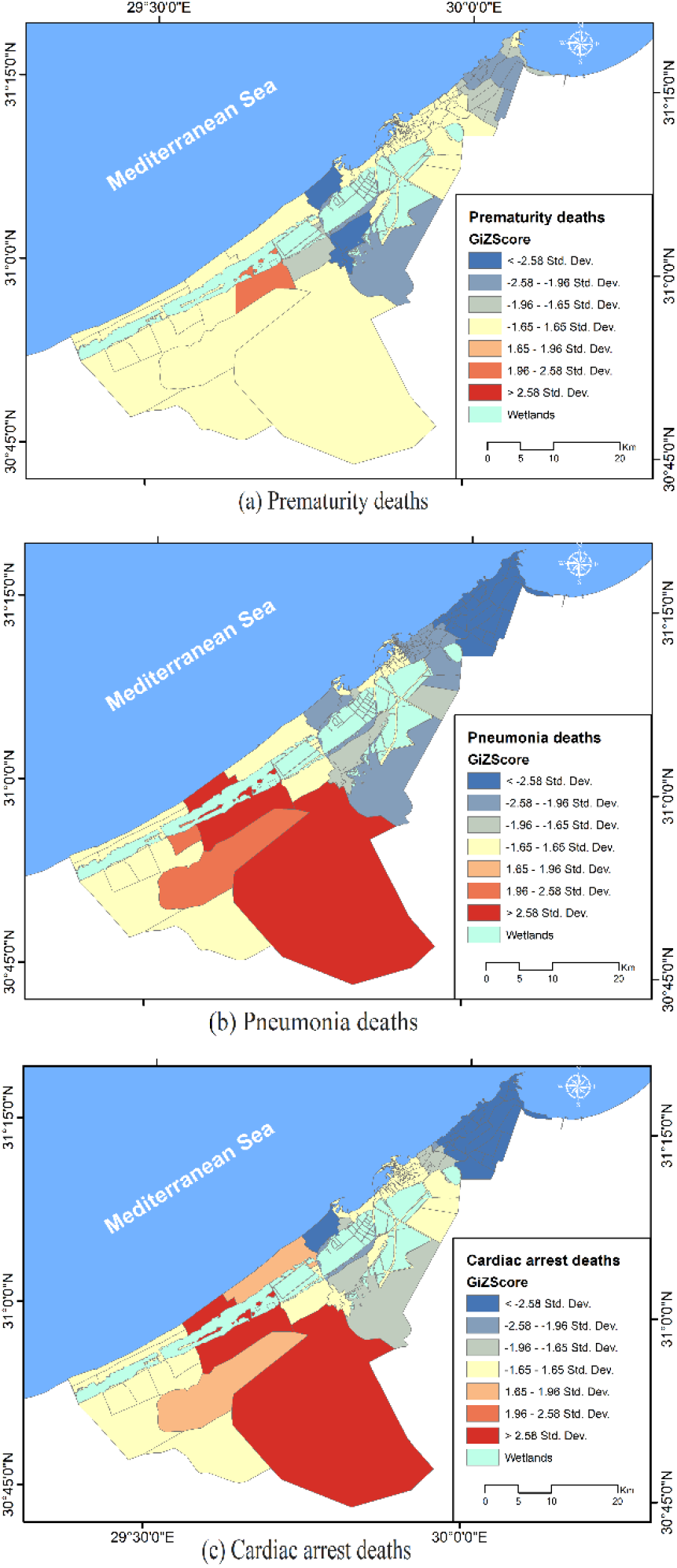
Hot and cold spots of main leading causes of under-five mortality.

## Cluster analysis (Two step cluster analysis algorithm)

The collected data for each locality varied to have some locality characteristics, illiteracy measures, and divorce measures in addition to the total recorded number of U5M. These data were subjected to two-step cluster analysis to classify the localities according to the observed differences in the collected data for each locality. Upon performing the analysis, the quality of the algorithm was good, having an average silhouette measure of 0.57. The algorithm classified the localities into 2 clusters, 17 localities (56.7%) in cluster 1 and 13 localities in cluster 2. The most important classification variable was the university grade for females whose average was 3.56 in the cluster 1 localities versus 22.98 in cluster 2 localities. The variables indicating illiteracy was most important, followed by the divorce variables and the locality characteristics’ variables. **Table** 3 Generally, localities in cluster 1 had higher rates of illiteracy and divorce along with a lower quality of the locality characteristics. It is worth noting that all hot spots of U5M were classified to cluster 1, indicating an interaction between some of the classification variables with the recorded U5M. In other words, all of the four hot spot localities were classified to cluster 1 (17 localities in total), it might be a warning sign for the other 13 localities in cluster 1 that they have a higher tendency for being hot spots of U5M compared to cluster 2 localities.

### PLS-SEM model

Upon performing the analysis, the loadings of the variables on the first latent variable “locality characteristics” indicated higher quality of locality characteristics, the loadings on the latent variable “divorce” indicated higher divorce rate, the loadings on latent variable “illiteracy” indicated higher illiteracy and lower university grade, and the loadings on the latent variable “mortality” indicated higher number of recorded mortalities. Illiteracy to mortality path and the divorce to mortality paths were statistically significant (*P*= 0.014 and 0.017, respectively). Locality characteristics explained about 43% of the divorce variance and about 68% of the illiteracy variance among the tested localities. The locality characteristics, divorce and illiteracy explained about 30% of the variance of the recorded U5M among the investigated localities. The divorce had the higher impact on mortality (path coefficient = 2.194) compared to illiteracy (path coefficient = 1.285) and locality characteristics (path coefficient = 0.533). **Figure 7**

**Figure 7:**
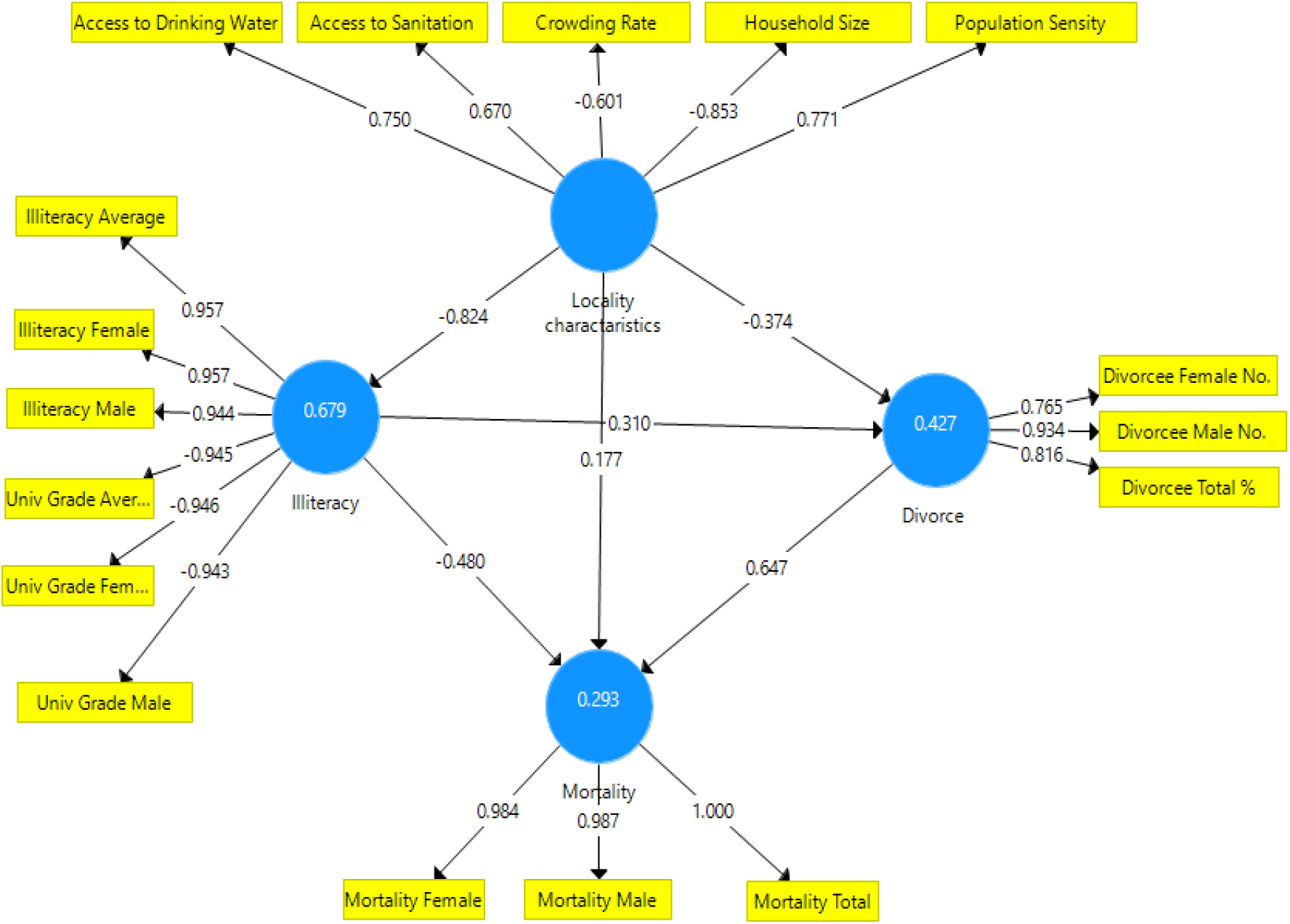
PLS-SEM model of the predictors of under-five mortality in Alexandria, Egypt

## Discussion

This study highlighted the tackled the geospatial distribution of U5M Alexandria Egypt. Pattern of U5M was not evenly distributed across the governorate. We identified hot spots in the western parts of Alexandria including four localities: El Hawaria, Bahig, Hamlis and Ketaa Maryiut. The main identified the main causes of U5M were prematurity, pneumonia, cardiac arrest, congenital malformation, and childhood cardiovascular diseases and determinants of U5M. In addition, this study addressed the main causes of deaths among different age groups (neonates, post neonates, and childhood) and mapped the most frequently reported three causes of U5M in Alexandria.

In this paper, about 41.3% (1207/2924) of U5M in Alexandria took place during the post-neonatal, and childhood period, this means that the majority of U5M takes place during the first year of life. It is worth mentioning that the trend of U5M rate in Egypt showed a remarkable decline during the last three decades, the number of U5M decrease from 86/10^3^ live-birth in 1990 to 20.28/10^3^ live-birth in 2019. Despite this decline in U5M, infant to U5M increased from 74.5% (63.7/85.5) in 1990 to 88.8% (19.86 /22.57) in 2016. Similarly, neonatal mortality to infant mortality increased from 53.1% (33.82/63.7) to 70.0%(13.91/19.86).^(21)^ Such an overall decline of U5M reflects the achieved improvement in health services provided by the Ministry of Health and Population including increase vaccination coverage, and maternal and health care services. Generally, the attained decline in U5M was more noticed among children than neonates. In fact, this uneven decline in mortality may be due to improvement in control and preventive strategies targeting mainly manageable and infectious diseases like pneumonia and diarrhea that may be more predominant in children. For instance, UNICEF report published in 2015 demonstrated that the proportion of U5M complaining of diarrhea decreased from 18.4% to 8.5%, and 14.0% in 2005, 2008, and 2014 respectively, more than 53% of these children receive health services. Similarly, 68.1% of children with pneumonia received health care, about 62.8% of them had antibiotics.^(22)^ On the contrary, despite achieving great reduction in neonatal mortality, many causes of death at this age may be more difficult to be managed or incompatible with life. For instance, congenital malformation and cardiac anomalies that may be inoperable or life-threatening. ^(23)^

In this research, the proportion of male deaths (53.2%) was significantly higher that female deaths (46.8%). This significant difference was evident during the neonatal period, while no differences were observed in the post neonatal and childhood period. Similarly, the Egyptian Demographic Health Survey conducted in 2008 reported that male deaths represented 52.5% of the U5M. ^(24)^ In the same line, UNICEF reported that U5MR was higher among males than females (21 vs 19 /10^3^ live births) ^(25)^ We can conclude that the sex related pattern of U5M did not change during the last few decade.

One striking revelation from the maps was that fatalities were not symmetrically distributed all over the study setting. Using GIS-based spatial analysis we identified that hotspots of U5M are in the western part of Alexandria governorate (El Hawaria, Bahig, Hamlis and Ketaa Maryiut). Of note, these localities are characterized by poor access to health services and low population sociodemographic conditions. Low sociodemographic conditions at the individual, household, or at the community levels are strong predictors of U5M as they affect the level of maternal education, housing condition, nutritional status, environmental pollution, family income, and exposure to injuries.^(26-28)^ In the same way, Malderen et al, ^(29)^ reported that U5M in Sub-Saharan Africa was strongly affected by socioeconomic status like child sex, maternal education, household wealth, and residency. Similar GIS-based study was conducted by, Anafcheh et al, ^(30)^ in Iran to determine the spatial distribution of U5M in Khuzestan province. They concluded that there was a great disparity of U5M a cross the province. Maternal factors like age, type of delivery, and birth intervals were significant predictors of U5M, however, mother level of education was not a predictor of U5M.

After clustering of the study localities, we identified the main characteristics of each cluster. Cluster 1 which contained the hot spots had a higher prevalence of illiterate females, divorced population, poorer locality characteristics in term of water supply and sanitary disposal, crowding index, and lower proportion of university graduated females. Literature proved that U5M is generally inversely related to mother’s educational level, with children born to women who never attended school being more than twice as likely to die by the fifth birthday as children born to mothers with secondary or higher education. Births to mothers in the highest wealth quintile are nearly three times as likely to survive to their fifth birthday as children born to mothers in the lowest quintile.^(24)^ The alarming finding was that 9 localities that were classified as cold spots were grouped as cluster 1. This means that soon or later these localities can convert to hot spots unless corrective actions are considered.

In this research, the most common identified causes of death among under-five were prematurity (28.32%), pneumonia (11.01%), cardiac arrest (10.57%), congenital malformation (9.95%), childhood cardiovascular diseases (9.20%), and septic shock (7.66%). In the same vein, World Health Organization reported that the most frequently reported causes of U5M were prematurity (29%), pneumonia including neonatal pneumonia (10.8%), and congenital malformation (11.6%), and birth asphyxia (9.8%)^(31)^. This means that infectious diseases remain the main killer of children in Egypt. Generally, these findings would effectively help stakeholders and policymakers in developing and implementation of more effective preventive and control measures, which can, finally, contribute to more robust reduction in U5M.

Points of strength: this study is one of the first few worldwide published studies that addressed the geospatial distribution of U5M. Furthermore, the large sample size of deaths included in the analysis and sampling technique greatly affect the external validity of the study outcome. Limitations of the study: this study was conducted within 1.5 years only; a study of longer duration may show different causes of U5M. Moreover, the data was collected from health offices where the full profile of deceased child was not available. Secondly, the recorded causes in death certificate were based on clinical assessment of the attending physicians. Autopsies were not performed on any of the deceased children, and there were no post-mortem records that could be used to verify the correctness of the causes of deaths recorded in the death certificate. Finally, this study was a retrospective cross-sectional study from records; this may affect cause-effect relationship due to vulnerable to bias.

## Conclusion

Spatial analyses of U5M clustering ate local level is essential for effective health policy intervention as it can highlight and explain intra-locality variations appropriately. In this respect. GIS can play an essential role in reducing U5M through delineating the hot spots of U5M and exploring various potential sociodemographic conditions at locality level. In this research, we identified four hot spots in Alexandria. There are 9 cold pots clusters that had similar characteristics to hotspots, and they are at risk to become hotspots. The key determinants of U5M were maternal education, divorce rates, and locality characteristics like household size, water supply, and access to sanitation. In future, effective management of these determinants can effectively reduce the observed disparity in U5M.

## Data Availability

Data available upon request

## Acknowledgment

Many thanks to archivist who provided us with all required data about the deceased children. We would like to acknowledge Prof Ahmed Mandil, and Dr Amr Rahal for revising this manuscript.

## Data availability

Data is available upon request by mailing the last author.

## Author Contribution

**Mahmoud Hassan**: Data cleaning, GIS analysis, and writing the manuscript.

**Ahmed Ramadan:** Statistical analysis, Writing manuscript.

**Ramy Ghazy**: Grant’s holder, field supervisor, data cleaning, statistical analysis, and writing manuscript. The rest of the team: data collection, and entry.

